# Contribution of quality improvement initiative in strengthening laboratory services in Primary Healthcare Facilities in Tanzania: findings from 2017/2018 Star Rating Assessment

**DOI:** 10.1101/2022.12.20.22283751

**Authors:** Erick Kinyenje, Ruth R. Ngowi, Yohanes S. Msigwa, Joseph C. Hokororo, Talhiya A. Yahya, Chrisogone C. German, Akili Mawazo, Mohamed A. Mohamed, Omary A. Nassoro, Mbwana M. Degeh, Radenta P. Bahegwa, Laura E. Marandu, Syabo M. Mwaisengela, Lutengano W. Mwanginde, Robert Makala, Eliudi S. Eliakimu

## Abstract

**Background:** Accurate and reliable diagnosis is the cornerstone of disease management and control. A reliable and properly organized laboratory system not only generates information critical to individual case management but also to disease surveillance, control, and outbreak management. This study presents the status of quality of laboratory services in Tanzania after a nationwide quality assessment, Star Rating Assessment (SRA) of PHC facilities conducted in 2017/18.

**Methods:** This was a cross-sectional study using secondary data from SRA dataset. Eight indicators were used to measure quality of laboratory services; of which together with facility characteristics are considered independent variables. Dependent variable is the general scores for laboratory services during SRA. Firstly, proportions of facilities for the indicators was calculated. Then, multiple linear regression was employed to determine impact of each variable on quality of laboratory services. P*-*value of < 0.05 was considered significant.

**Results:** Approximately one-quarter of 6,663 PHC facilities included, i.e., 1,773(26.6%) had appropriate staffing level for qualified health laboratory personnel. The situation was better in private facilities compared to public facilities (63% vs 19%, p<0.001); and in urban-based facilities compared to rural-based facilities (62% vs 16%, p<0.001). None of the indicators was complied with at least half of the facilities. Three indicators were the strongest positive predictor of laboratory quality scores: having a laboratory safety system (*Beta = 3*.*403*), availability of essential laboratory tests with SOPs available and adhered (*Beta = 2*.*739*), and appropriate staffing level for laboratory personnel (*Beta = 1*.*498*). The scores were likely to be low if the facility was a dispensary (*Beta = -1*.*325*), located in a rural area (*Beta = -0*.*068*) or publicly owned (*Beta = -0*.*048*).

**Conclusion:** There is a critical shortage of qualified laboratory personnel in PHC facilities, especially in public facilities that are based in rural areas. There is a need to further strengthen laboratory services in PHC facilities to ensure quality of laboratory test results, since none of the indicators was complied with at least half of the facilities.

## INTRODUCTION

In the past two decades (2000 – 2020), Tanzania has made significant efforts to strengthen its health system as part of the then health sector reforms that started in 1994; [1] coupled with interventions for health system strengthening following the World Health Organization (WHO) - World Health Report of 2000 [2] and the “*WHO-everybody’s business - strengthening health systems to improve health outcomes: framework for action*” of 2007[3]. The efforts included construction and rehabilitation of health infrastructure at all levels of health services delivery, procurement of equipment, and capacity building of health personnel. The efforts have enabled improvement in diagnostic availability in the primary health care (PHC) facilities. For example, a recent analysis has shown that diagnostic availability in dispensaries and health centres in Tanzania, have increased by 6-percentage points and 7.8-percentage points respectively from 2006 to 2014 although there are variations across the country[4].

Specific efforts that aimed at strengthening and improving the quality of laboratory services provided included the following: (i) harmonization and standardization of laboratory equipment in various levels of health services delivery including PHC facilities (mainly district hospitals and health centers) in order to address the challenges on the areas of procurement of reagents, maintenance of equipment, and quality assurance; [5 6] (ii) establishment of the National Health Laboratory Quality Assurance and Training Centre since 2008 with “*the purpose of improving quality of laboratory services all over the country*” currently referred to National Public Health Laboratory; [7] (iii) development and implementation of the “*National Health Laboratory Strategic Plan 2009 – 2015*”; [8] (iv) implementation of the World Health Organization African Region Stepwise Laboratory Improvement Process Towards Accreditation (WHO AFRO SLIPTA) in 2009 and implementation of the laboratory training program (Strengthening Laboratory Management Towards Accreditation (SLMTA)), in 2010; [9] and (v) development of the Point of Care Testing (POCT) Certification Framework in October 2017 aiming at improving the quality of HIV testing services in health facilities. [10] In Africa, Laboratory services especially in the area of HIV have improved significantly with potential for supporting country systems in future global health threats [11] since the Maputo Declaration of 2008 [12].

Importance of a strong PHC system has been further demonstrated by the ongoing coronavirus disease of 2019 (COVID-19) pandemic in which its effects have shown clearly the need for a strong PHC as a means to uphold equity and strengthen capacity to respond to emergencies[13]. Laboratory services in PHC facilities have been shown to enhance performance of PHC facilities [14]. Also, the framework produced by the Lancet Global Health Commission for High Quality Health Systems in Sustainable Development Goals Era, shows that improving laboratory services in PHC facilities is an essential element for ensuring “*competent care and systems*” which is one of the components of processes of care in the framework[15].

Access to laboratory services at district level in Tanzania has been shown to be inequitable in the sense that in some areas, people have longer travel time to access a nearby facility for the services[16]. An analysis of data from 10 countries including Tanzania, has shown that only 199 (2%) of the facilities investigated in those countries had all of the diagnostics services[17]. Also, laboratories in PHC facilities have been reported to face a number of challenges in supporting care and treatment services for HIV services which include “*number of qualified personnel, staff training on the national guidelines, laboratory diagnostic tools and coordination*” [18]. In a study conducted in 2014, basic diagnostic equipment for HIV and diabetes were observed more frequently in hospitals than in health centres and dispensaries[19]. Strengthening laboratory services in PHC facilities will help to capacitate Tanzanian PHC system to tackle the challenge of Non-Communicable Diseases (NCDs) [20]. In Ghana, improvement of POCT diagnostic services in PHC facilities have been recommended as part of improving maternal health services [21].

The Government of Tanzania embarked on an initiative called “*Big Results Now*” in health sector since 2014/2015 with several initiatives among them being performance management of PHC facilities through implementation of Star Rating Assessment (SRA). The SRA was implemented in Tanzania mainland in three period of time: baseline assessment was conducted in the fiscal year 2015/16 all 26 regions; second assessment was conducted in fiscal year 2017/18 in all 26 regions; and third assessment was conducted in fiscal year 2021/22 in 10 regions. The assessments used a set of tools for dispensary, health centre and hospitals at council level, each with 12 service areas that were assessed [22 23]. Service area number 12 is on clinical support services in which one of the sub - area is laboratory services [24]. Therefore, the purpose of this paper is to describe the status of laboratory services in the PHC facilities during the 2017/18 second assessment in order to recommend on measures appropriate for further strengthening of the services towards achievement of the universal health coverage (UHC). Findings of the study will also contribute in understanding the situation of PHC delivery in a way that will be in line with the new health system performance assessment framework to inform policy decisions [25].

The specific objectives of this study are to: determine proportion of PHC Facilities that comply with staffing level for qualified laboratory personnel; determine the proportion of PHC facilities that comply with other indicators for measuring the quality of laboratory services; and identify the predictors for high scores in indicators that measure the quality of laboratory services among PHC facilities.

## METHODS

### Study design

This was a cross-sectional study using secondary data that were collected from all PHC facilities during star rating second assessment in 2017/18. The data are available in the national data platform, i.e., DHIS2.

### Study population

The healthcare facilities in Tanzania can be grouped into two major categories; PHC facilities and Referral Healthcare Facilities (RHFs). RHFs are referral points for all PHC facilities, these are hospitals that are distributed in all 26 regions of the country. Within each region, there are a number of PHC facilities ranging from dispensaries, health centres to hospitals at the council level, in that order of expertise. Dispensaries provide exclusively outpatients’ services to about 10,000 populations. Health centres are designated as referral points for dispensaries because they offer a broader range of services including inpatient services and Comprehensive Emergency Obstetric and Newborn Care (CEmONC) to about 50,000 populations. A hospital at the council level (i.e., level 1 hospital) serves about 250,000 population and receives referrals from the low levels [26].

Additional facility characteristics such as the place where the facility is located (urban or rural), and ownership status (private or public) may significantly affect access, management and quality of services delivered. Between 2017 and 2018; SRA was conducted to 7,289 PHC facilities that are unrestrictedly distributed all over the country. PHC facilities constitute about 95% of all healthcare facilities in Tanzania.

#### Inclusion criteria

This study includes all PHC facilities that completed successfully SRA conducted in 2017/18.

#### Exclusion criteria

Any facility whose scores on quality of laboratory services were not found in the SRA database was excluded from the study.

### Star Rating Assessment database

SRA database is a part of DHIS2 platform that is managed Ministry of Health, Health Quality Assurance Unit (HQAU). Paper based tools were used to collect baseline data at 2015/16 assessment while data were collected electronically through DHIS2 during the second and third assessment held in 2017/18 2021/22. Since the dataset for baseline were not reliable; we used second data assessment dataset for this study. The dataset is further divided in 12 quality assessment areas as per SRA tool. Area number 12 is namely clinical support services, one of the support service was laboratory services. The section contains score data on eight indicators of quality of laboratory services, as presented in Table 1.

**Table 1:**
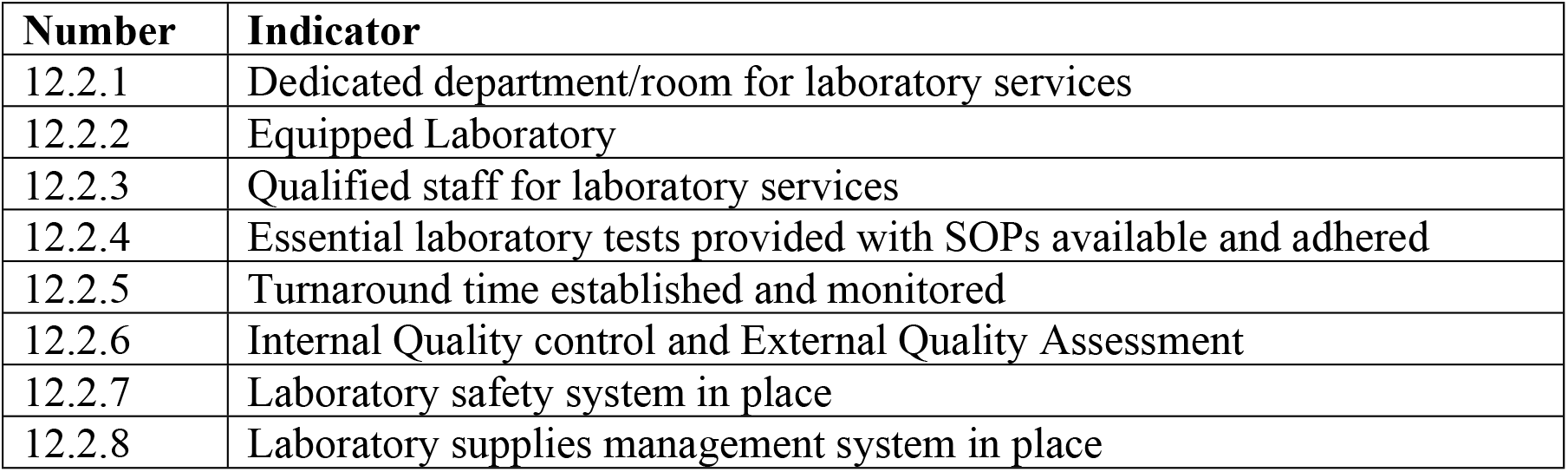
Service Area 12 (Clinical supportive services): sub-Area 12.2 on Laboratory Services.

### Data collection and variables

#### Independent variables and how were collected

The eight indicators of the quality of laboratory services (Table 1) were considered independent variables for this study. The indicator was scored “1” whenever the facility had complied with staffing level for qualified laboratory personnel, otherwise “0”. As per Tanzanian “Staffing Levels for the Ministry of Health and Social Welfare Departments, Health Service Facilities, Health Training Institutions and Agencies of 2014-2019” [27]; the dispensary was required to have at least one Laboratory Assistant, a health centre to have one Laboratory Technologist and one Assistant Laboratory Technologist. The hospital was to have minimum of five personnel such as two Assistant Laboratory Technologist and three Laboratory Technologists.

The facilities that had a dedicated department or room for laboratory services scored “1” for the indicator number 2, otherwise “0”. A dedicated room should have been equipped with running water and the basin, plus suitable worktop. Furthermore, a facility needed to have a laboratory equipment with the necessary equipment to score “1” for indicator number 3. Despite many of equipment and or supplies requirement for PHC [6] few of them were selected and included during SRA assessment. These include Haemoglobinometer, POC glucometer, tubes for collection of blood, and containers for collection of urine specimens for dispensaries, while microscope and centrifuge were added for health centres and hospitals.

Indicator 4 determines facilities with essential laboratory tests with SOPs available and adhered. Three important sub-indicators were summed up to make this indicator: availability of a full package of recommended tests; availability of Standard Operating Procedures (SOPs) for the tests; and whether the SOPs were adhered for at least three randomly chosen tests. Test menu for laboratory services identified many test for PHC[28] however in SRA few test were selected such as a test for malaria, Hb, urinalysis, UPT, blood glucose level for dispensaries and addition of HIV test to health centres and hospitals. Availability of tests was considered “1” if all of the mentioned test were conducted within a week before the assessment.

The Facility scored indicator 5 (turnaround time established and monitored) if had established turnaround time for tests performed and it is monitored [28 29]. Additionally, the facility scored “1” for indicator 6 if the laboratory had performed internal quality control and participated in external quality assessment (EQA). To score this indicator; the facility must had performed internal quality control and participated in external quality assessment (EQA) previous quarter year, results documented, corrective and preventive actions taken.

Indicator 7 scored “1” if the laboratory had a safety system in place. Safety systems for all facilities included the presence of safety-related procedures such as availability and adherence of: laboratory safety rules, IPC SOPs that include SOPs for post-exposure prophylaxis and healthcare waste management [30 31]. An additional requirement for health centres and hospitals to score the indicator was whether these facilities had a refrigerator for provision of blood bank services.

Presence of Laboratory supplies management system was the last indicator variable. Facility scored “1” if had documented the incoming and outgoing stock in the register correctly. Three laboratory reagents and or consumable supplies were selected and their records tracked to determine if were adequately updated.

#### Dependent variable

Each facility was assigned an overall quality score for laboratory services during assessment. The score was regarded as dependent variable of the study that has been the result of the sum of individual scores from each question.

#### Statistical analysis

Data were checked for quality before analysis. Stata version 15 was used for data input and statistical calculations. The prevalence of eight independent variables of the quality of laboratory services among PHC facilities was analyzed by the Chi-square test. Multiple linear regression analyses were conducted using the laboratory quality score as the dependent variable. For all analyses, a p-value of < 0.05 was considered significant.

## RESULTS

### Characteristics of PHC facilities involved in the study

A total of 6,663 PHC facilities were included in this study, whereby 5,485 (82.3%) were publicly owned and 1,178(17.7%) were privately owned. The majority of the facilities were the dispensaries 5,721(85.9%) followed by health centres 732 (11.0%) and level 1 hospitals 210(3.2%). About one-quarter of the facilities were from urban areas 1,528(22.9%) and the rest were from rural areas 5,135(77.1%).

### PHC Facilities that comply with staffing level for laboratory services

About one-quarter of the PHC facilities, i.e., 1,773(26.6%) had complied with recommended number of qualified health laboratory personnel (Table 2). As presented in Table 3; the compliance was significantly highest at hospitals (40%), followed by health centres (31%) and then dispensaries (26%) (p<0.001).

**Table 2:** Availability of each allocated qualified laboratory personnel among Tanzanian PHC facilities 2. 1.2.

| Variable | Staffing | Yes |  | No |  | PHC facilities complying with staffing level requirements |  |
| --- | --- | --- | --- | --- | --- | --- | --- |
| PHC facility level | Type of Laboratory Personnel | n | % | n | % | n | % |
| Dispensaries (n=5,721) | Laboratory Assistant (1) | 1,464 | 26% | 4,257 | 74% | 1,464 | 26% |
| Health Centres (n=732) | Laboratory Technologist (1) | 263 | 36% | 469 | 64% | 225 | 31% |
|  | Assistant Laboratory Technologist (1) | 559 | 76% | 173 | 24% |  |  |
| Hospitals (n=210) | Laboratory technologist (3) | 94 | 45% | 116 | 55% | 84 | 40% |
|  | Assistant lab technologist (2) | 158 | 75% | 52 | 25% |  |  |
| **All PHC facilities** |  |  |  |  |  | 1,773 | 26.6% |

**Table 3:** PHC facilities with appropriate staff (laboratory personnel) for provision of laboratory services on the day of assessment (N=6,663)

| Variable | Yes |  | No |  | P-Value (chi sq test) |
| --- | --- | --- | --- | --- | --- |
| PHC facility level | n | % | n | % | <0.001 |
| Dispensaries | 1464 | 26 | 4257 | 74 |  |
| Health Centres | 225 | 31 | 507 | 69 |  |
| Hospitals | 84 | 40 | 126 | 60 |  |
| PHC facility ownership |  |  |  |  | <0.001 |
| Public | 1,036 | 19 | 4,449 | 81 |  |
| Private | 737 | 63 | 441 | 37 |  |
| PHC facility location |  |  |  |  | <0.001 |
| Rural | 827 | 16 | 4,308 | 84 |  |
| Urban | 946 | 62 | 582 | 38 |  |

The compliance to staffing level was significantly higher in private facilities compared to public facilities 63% vs 19% (p<0.001). Likewise, it was better among urban-based facilities compared to rural based facilities (p<0.001) (Table 3).

### The proportion of PHCs that comply with indicators that measure the quality of laboratory services

As presented in Table 4, the indicator that performed highest (45.1%) by all-level PHC facilities was “facilities with a dedicated department/room for laboratory services” while most of the facilities performed poorly in implementation of Internal Quality control and External Quality Assessment (22.2%). Less than half of the facilities at the hospital-level and health centre-level had refrigerator for blood transfusion services.

**Table 4:**
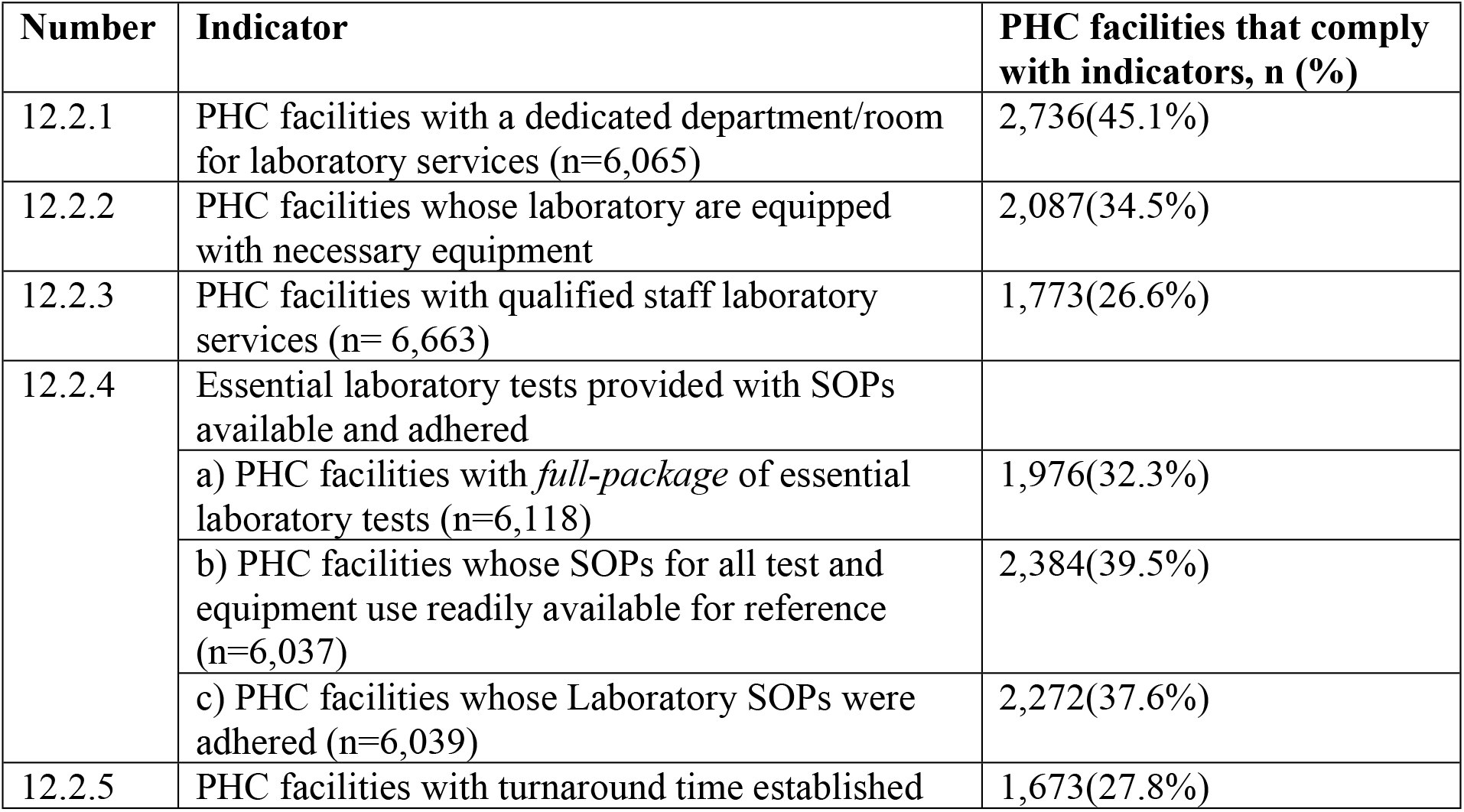
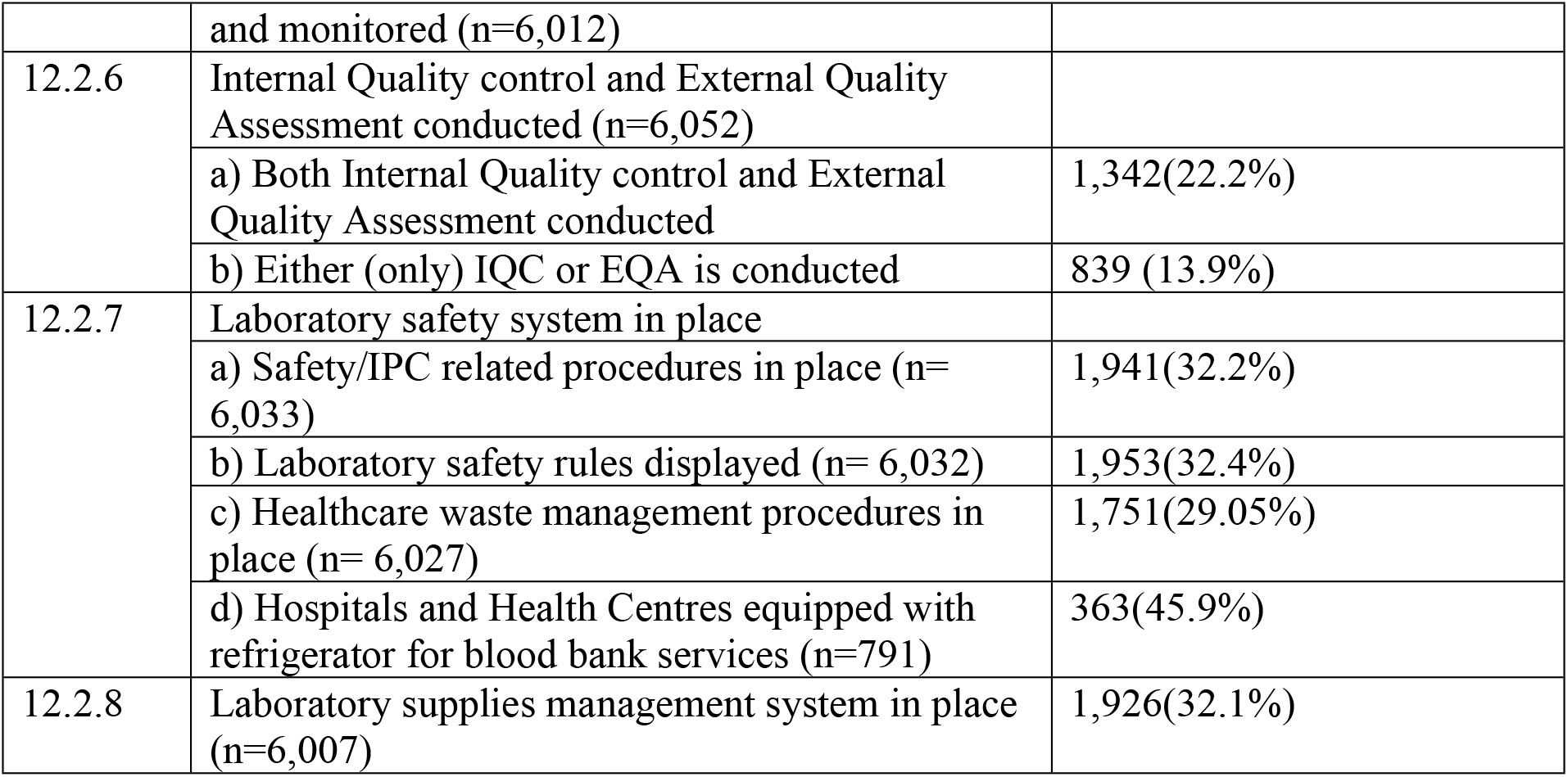
Proportion of PHC facilities that comply with indicators that measure the quality of laboratory services (N=6,663)

### Predictors for high scores in indicators that measure the quality of laboratory services among PHC facilities

The findings from multiple linear regression analysis suggest that indicators that measure the quality of laboratory services and the characteristics of PHC facilities had a significant effect in predicting scores in the quality of laboratory services among PHC facilities. The variables explain 99.1% of the variance in laboratory service scores in the model.

Among these predictors, having the laboratory safety system in place (*Beta = 3*.*355*) was found the strongest positive predictor followed by the availability of essential laboratory tests with SOPs available and adhered (*Beta = 2*.*857*), and complying with standards for qualified laboratory personnel (*Beta = 1*.*176*) in defining laboratory quality scores positively. On the contrary, the following facility characteristics were negatively defined by the scores in laboratory services; facilities at dispensary level (*Beta = -1*.*947*), facilities that are located at the rural area (*Beta = -0*.*033*) and facilities that are owned by public organizations (*Beta = - 0*.*061*). The details are shown in Table 5.

**Table 5:**
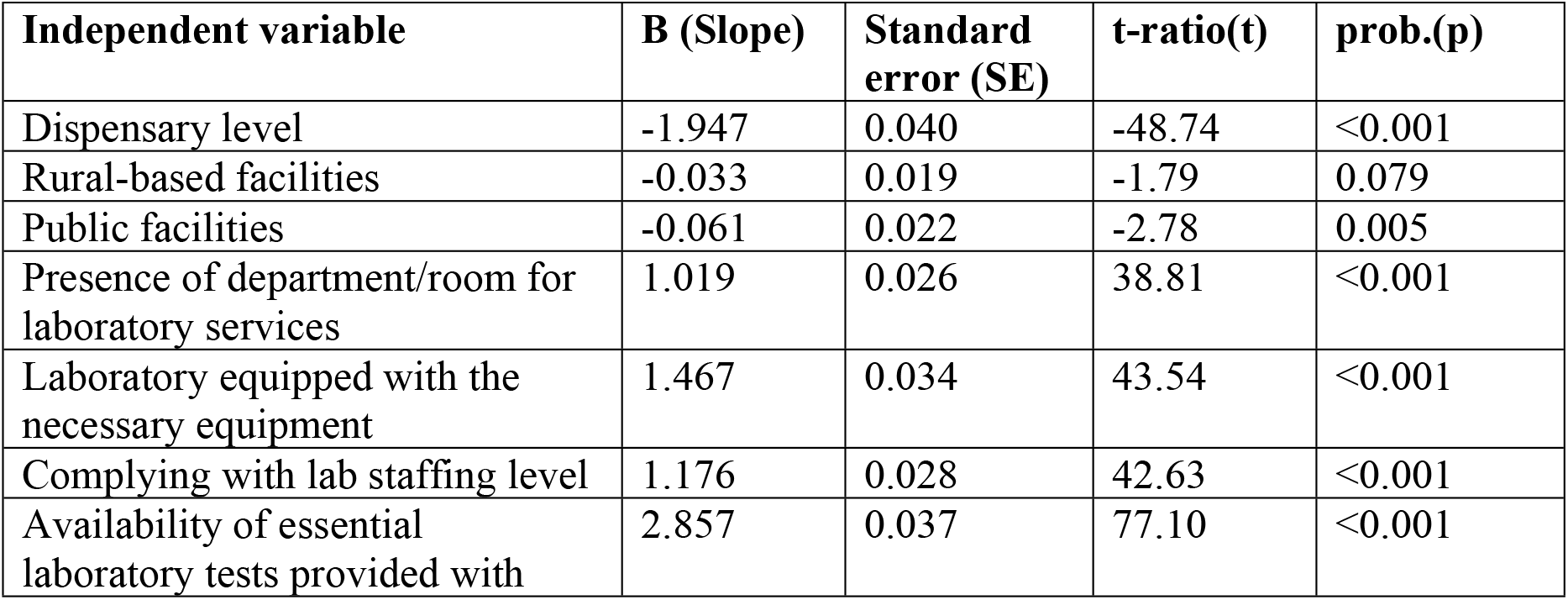
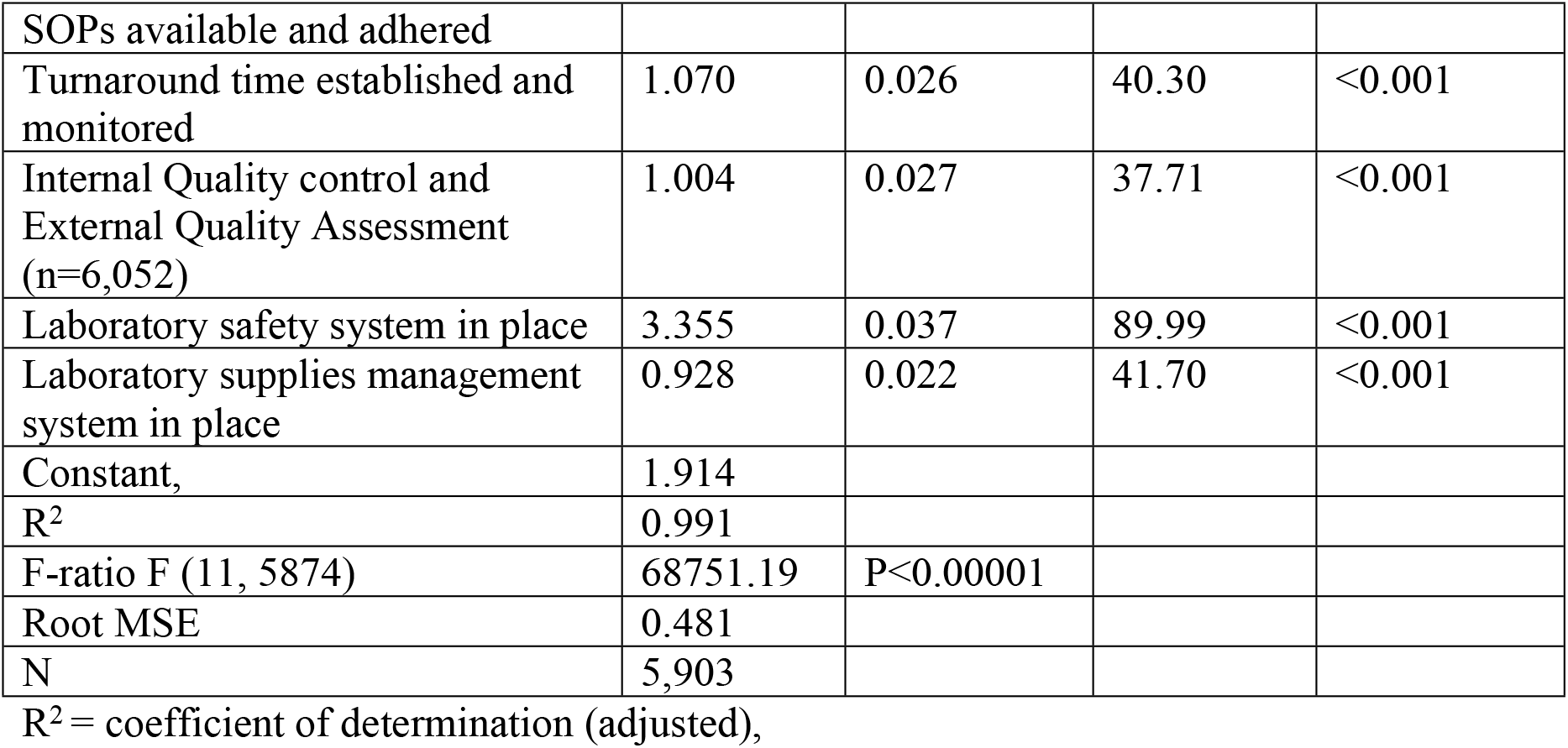
Predictors of PHC facility scores for laboratory quality services, 2017-2018.

## DISCUSSION

The study has shown that the PHC facilities in Tanzania had low level of compliance with all the laboratory indicators used in the SRA as per the 2017/2018 dataset of the results. The low performance in laboratory quality indicators has also been reported in other studies. For example, a study in Ethiopia also has reported that PHC facilities had inadequate access to essential laboratory tests; [32] and another study also in Ethiopia found that unavailability of laboratory tests in government owned health centres in southern Ethiopia affected outpatients service utilization causing most diagnoses to be based on clinical findings [33]. A study in four regions in Tanzania found that few dispensaries were providing diagnostic services for HIV. Also, studied facilities (hospitals, health centres and dispensaries) had inadequate staff, lack of maintenance of the equipment, and that few facilities conduct Internal Quality control and few participated in External Quality Assessment programmes [34]. Also, in Kenya inadequate availability of tests for infectious diseases and other diseases has been shown to affect the readiness of PHC facilities to support the country in implementation of its programme towards attainment of the UHC target [35]. Odjidja and colleagues have reported that availability of diagnostics for Malaria, Tuberculosis and HIV is one of the predictors for pregnant women to receive integrated care for all the three diseases [36].

### Policy implications

Our study has two important policy implications. First, the results provide a status of a countrywide laboratory services quality and capacity in the PHC facilities which will help in the ongoing efforts to strengthen performance of PHC facilities, as well as in the processes for reimagining PHC services post COVID-19 [37]. Secondly, based on these findings the Ministry need to strengthen further the laboratory services in order to be able to support some disease specific interventions. For instance, in a study conducted in three districts (Uyui, Geita and Ukerewe) in north-western Tanzania involving 80 PHC facilities to assess capacity for integrating schistosomiasis control activities found that only 33.8 % (27/80) of the PHC facilities had laboratory services for diagnosis of schistosomiasis [38].

Therefore, the MoH in collaboration with PORALG and health sector stakeholders need to further strengthen laboratory services in all PHC facilities, as part of improving and maintaining the quality of laboratory services provided [39]. In doing so, the Ministry will be harnessing its work on the Sustainable Development Goal 3 (*ensure healthy lives and promote well-being for all at all ages*); target 3. 8: “*achieve universal health coverage, including financial risk protection, access to quality essential health-care services and access to ……*.”, [40] since diagnostic services are an essential ingredient of the UHC target [41].

## CONCLUSION

There is a critical shortage of qualified laboratory personnel needed to improve the quality of laboratory services in Tanzania, especially in public facilities that are based in rural areas. There is a need to further strengthen laboratory services in PHC facilities to ensure quality of laboratory test results generated since even in the laboratories that were found to be working, more than half did not conduct Internal Quality control and or participating in External Quality Assessment. The overall compliance with all laboratory indicators is inadequate (none was achieved by at least half of the facilities). Availability of: qualified laboratory personnel; equipped laboratory with a full range of laboratory tests and standard operating procedures adhered; laboratory safety systems are key in improving the quality of laboratory services.

## Data Availability

Data for this study can be requested from Principal Secretary of Ministry of Health through

